# Growth, infection, and humoral immunity in children who are HIV exposed and uninfected

**DOI:** 10.64898/2026.02.25.26347096

**Authors:** Romeo Djounda, Romuald Ngamaleu, Honore Awanakam, Moritz Schmiedeberg, Kevine Tchamda, Martial Tsague, Eva Gutenkunst, Jude Bigoga, Rose Leke, Charles Kouanfack, Micheal Besong, Krystelle Nganou-Makamdop, Forgu Esemu Livo

**Author notes:** Corresponding author: Forgu Esemu Livo.

## Abstract

**Background:** Children who are HIV-exposed uninfected (HEU) show greater morbidity and mortality than HIV-unexposed children (HUU). In this study we investigate sex differences in growth, infection rates and antibody response among HEU and HUU infants.

**Methods:** The study enrolled 107 pregnant women with HIV and 103 pregnant women without HIV with follow-up of their infants from birth to 12 months of age. Study measures assessed included growth parameters, the prevalence of children with overt disease symptoms as reported by the mother, PCR-based assessment of infections (cytomegalovirus (CMV), respiratory syncytial virus (RSV), rhinovirus, influenza A & B, rotavirus and malaria) as well as antibody profile to CMV, RSV and enterovirus infections.

**Results:** Compared to male HUU, male HEU infants had lower Height-for-age-z-scores (β -0.75; P=0.047) in mixed-effect model accounting for age. Additionally, they showed transiently lower Weight-for-age-z-scores at 3 months (1.07 vs 0.05, P=0.04), with higher risk of rhinorrhea (RR=2.29, P=0.02) and lower enterovirus titers at birth (P=0.0066). Female HEU showed transiently higher stunting at 6 months (0% vs 21%; P=0.01) and lower CMV viremia at 6 months, with elevated CMV antibody titers at 3 months (P=0.04) compared to female HUU. With prevalence ranging from 25%–61%, CMV and Rhinovirus infections were dominant in all groups. HEU and HUU exhibited similar antibody decay and acquisition patterns for CMV, RSV, and Enterovirus across both sexes.

**Conclusion:** HEU infants show transient sex-based differences in growth, infection and immune profiles raising the relevance for considering sex as a key parameter to assess infant health.

## Background

The implementation of the prevention of mother to child transmission (PMTCT) has greatly averted the number of perinatal HIV infections. This has led to a steady increase in the population of children who are HIV-exposed but uninfected (HEU) as born to cART-treated women living with HIV(WLHIV). Currently there are over 16 million HEU worldwide with over 90% of them living in Sub Saharan Africa where the burden of HIV remains highest[1]. Despite being uninfected with HIV, high mortality and morbidity rates have been reported amongst the HEU when opposed to their HIV unexposed uninfected (HUU) peers[2–4] Compared to HUU, HEU have been shown to have greater infection rates, infection related hospitalization and prolonged hospitalization[5–9]. Moreover, several studies report high prevalence of stunting and wasting amongst the HEU infants during their first 2 years of life[10–13]. The reasons for these differences are multifactorial and poorly understood. It has been hypothesized that exposure to cART *in-utero* and intrauterine chronic viral infections such as HIV antigens and CMV, reduced duration of breastfeeding, reduced maternal transfer of antibodies during gestation and breastfeeding, and reduced subsequent immune modulation in HEU infants are possible mechanisms that underpin differences in HEU and HUU infants[14]. The importance of humoral immunity is crucial since the infants rely on acquired maternal antibodies during their first months of life while their immune system fully develops[15]. Any quantitative and qualitative abnormalities in maternal antibody transfer and ulterior dynamics may cause imbalances in immune response against common infections. Infections with malaria, enteroviruses, cytomegalovirus (CMV) and respiratory syncytial virus (RSV) are major contributors to child morbidity and mortality among infants in low and middle-income countries (LMIC). Evidence shows that HEU infants experience reduced transfer of maternal antibodies to malaria, RSV and Influenza B[16–18]. The latter added to exposure to cART and HIV *in-utero*, might enhance susceptibility to infections. Despite the clinical importance of these infections, their true burden is unknown, and host response is understudied in HEU infants. Most studies have focused on individual aspects of HEU’s health, describing infection burden to single infections or evaluating immune responses in isolation. In addition, sex differences in infection dynamics and immune response are increasingly recognized and studied, but not extensively in HEU[19]. To address this gap, we recorded growth parameters, self-reported disease symptoms, infection burden for CMV, malaria, rotavirus, rhinovirus, RSV, Influenza A and B, and antibody dynamics to CMV, RSV and Enterovirus over the first year of life, and performed sex stratified analysis in a longitudinal cohort of HEU and HUU. These aspects put together can give a holistic view of overall trends in morbidity in HEU compared to their HUU counterparts.

## Methodology

### Study site and participants

This study was a prospective cohort study conducted in Yaoundé, Cameroon, at the CASS Nkoldongo hospital from June 2022 to May 2024. Pregnant women living with and without HIV were recruited in the third trimester of pregnancy and were received during delivery for the recruitment and follow-up of their HEU and HUU until 12 months of age. Inclusion in the study was restricted to women that had uncomplicated pregnancies and were either HIV infected on cART or uninfected and were not planning to relocate in another city in the next two years. Aligned with the national immunization schedule, study visits occurred at 6, 10 and 14 weeks (3 months) then at 6, 9 and 12 months.

This study was approved by the Cameroon Baptist Convention Health Boards’ Institutional Review Board (IRB2022-05) and all participants provided written informed consent. Research was conducted in accordance with the ethical standards of the Helsinki Declaration.

### Anthropometric and clinical assessments

Data collected during each visit included anthropometric measures (weight, length/height, mid upper arm circumference and head circumference) and clinical parameters (maternally-reported history of infection, hospitalization and symptoms). Major symptoms reported were solely based on mother observations: fever (perceived rise in temperature either by touch or using a thermometer: axillary temperature above ≥ 38°C), cough (forceful exhalation of air, sometimes sounding dry, wet or backing), diarrhea (waterier and more frequent stool in 24H), rhinorrhea (drainage of liquid or mucus from the nasal passage), skin rash (appearance of red spots, patches or irritation on the skin) and vomiting (forceful spiting of stomach contents). Growth Z-scores; weight-for-age (WAZ) length/height for age (HAZ), head circumference-for-age (HCZ), body mass index for age (BAZ) and weight height-for-age (WHZ) were calculated according to World Health Organization (WHO) standards using the WHO Anthro survey Analyzer.

### Sample collection and processing

Venous blood samples were collected from mothers at recruitment and from infants at birth (cord), 14 weeks, 6, 9 and 12 months. Maternal and cord plasma was separated and stored at -80°C. At all infants timepoints, aliquots of 100ul whole blood were stored in 100ul DNA/RNA shield (DNA/RNA shield^TM^, ZYMO RESEARCH) and stored at -20°C. Infants buccal, Nasal and Rectal swabs were also collected at 14 weeks, 6, 9 and 12 months and were conserved in DNA/RNA shield diluted 1:1 and frozen at -20°C.

### Detection, of pathogen infections by RTPCR

Nucleic acids were extracted from buccal, nasal, rectal swabs and whole blood for the screening of CMV, RSV, Rhino virus, Influenza A and B (Infl A & B), Rota virus, and malaria using ZYMO_RESEARCH extraction kits following manufacturers procedures. To screen for CMV, DNA was extracted from buccal swabs using Quick-DNA/RNA^TM^ viral kit. Likewise, to screen for malaria, DNA was extracted from whole blood specimens using Quick-DNA^TM^ miniprep kit. To screen for RNA viruses (RSV, Rhinoviruses, Infl A & B and Rota viruses), RNA was extracted using the Quick-RNA^TM^ viral kit. Amplification was conducted by quantitative real-time PCR (qPCR) for DNA and reverse transcription qPCR (RT-qPCR) for RNA on the Bio-Rad CFX_Opus_96 Real-Time PCR platform. Plasmid standards were also used to generate ten-fold serial dilutions (10^1^–10^7^ copies/µL) for absolute quantification. CMV was co-amplified with Albumin, to allow for normalization of cell input. All reactions were run with specific primers and probes using Luna Universal Probe One-Step RT-qPCR Kit w/o ROX and Luna Universal Probe qPCR Master Mix **Supplementary table 1 and 2.** Viral loads below the lower limit of detection (10copies/µL) were arbitrarily attributed a viremia of 8 copies/µL. CMV viremia was normalized to that of Albumin (two copies/cell) and expressed as per 10^5^ cells using the ratio of CMV to albumin copy numbers.

### Antibody measurements

CMV, RSV and Enterovirus specific total IgG antibodies were measured by ELISA. High binding ELISA plates were coated with recombinant CMV gB, RSV cell culture extract and recombinant enterovirus antigen (from Serion_Immunologics) at concentration of 1µg/ml in carbonate buffer (15mM NaCO3, 35mM NaHCO3 in H2O pH 9.6) and stored overnight at RT. Plates were blocked with 5% skimmed milk in PBS-T20 (PBS and 0.05% Tween-20) for 2h at RT. Plasma samples were diluted 1:200 in 2% skimmed milk PBS-T20 and incubated for 2h at RT. Goat anti-human IgG-HRP (1:3000) was then added and incubated for 1h at RT. After every incubation step, the plate was washed 3 times with PBS-T20. Subsequently, plates were washed twice with PBS prior to adding ECL solution and immediate measurement of relative light units per second (RLU/s) usng a microplate luminometer (Molecular Devices).

### Statistical analysis

Data collected was analyzed using GraphPad prism 10.6.0. Gaussian distribution of data sets was tested and corresponding parametric and non-parametric tests were applied for the comparison of continuous data. Paired tests were used to assess differences within dependent datasets. For analysis of categorical data, fishers exact or chi square tests were used. To assess the independent effect of HIV exposure on growth parameter over time, mixed effect model regression analysis was fitted using STATA 15.0. Covariates included Socioeconomic Status (SES), maternal and gestational age. For model parsimony, covariates that did not influence growth trajectories were excluded from model (maternal age and gestational age). Correlation analysis of antibody titers was performed using R software (version 4.5.1). P<0.05 defined significance.

## Results

### Participant characteristics

A total of 210 pregnant women were recruited among which 107 were living with HIV on cART and 103 without HIV. Of these, 195 were retained at delivery with 196 live births (94 HUU and 103 HEU) after excluding perinatal deaths. After excluding postnatal deaths, 193 children participated in at least one of the 6 scheduled visits with a retention rate of 63.8% with no perinatal HIV recorded at 6 weeks routine HIV-1 PCR (**Figure 1**).

**Figure 1:**
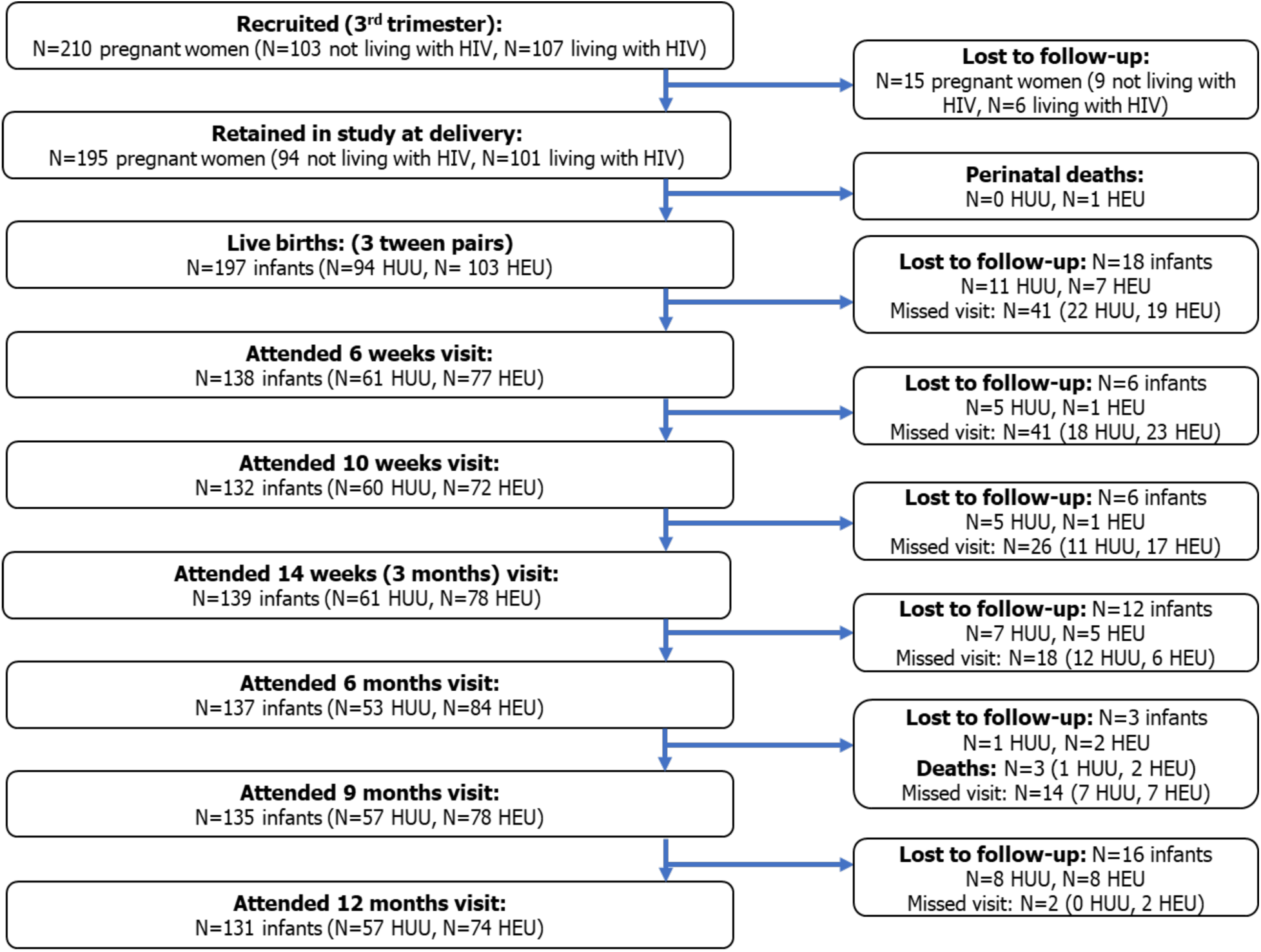
Flow diagram of included participants. Number of participants included in the study from recruitment of pregnant women through follow-up during the first 12 months of life. HEU: children who are HIV-exposed and uninfected; HUU: children who are HIV-unexposed and uninfected; HIV: human immunodeficiency virus.

When compared to pregnant Women living without HIV, women living with HIV were older (32 SD±5.7 vs 28 SD±5.4: P<0.0001) and multigravida. Women living with HIV were all on ART with most of them (76/107 [71%]) receiving the standard first-line ART regimen consisting of tenofovir_disoproxil_fumarate (TDF), lamivudine (3TC), and efavirenz (EFV) while 31/107 (29%) were on other combinations. The median duration on ART was 70 months (IQR = 23.3 – 103.8) **Table 1**.

**Table 1:** Maternal and child baseline characteristics.

| Parameter | Overall (N = 210) | Women not living with HIV or HUU (N = 103) | Women living with HIV or HEU (N = 107) | P value |
| --- | --- | --- | --- | --- |
| **Sociodemographic parameters** |  |  |  |  |
| Age in years, mean (SD) <sup>b</sup> | 30 (±5.7) | 28 (±5.4) | 32 (±5.7) | <0.0001 |
| Weight (kg), median (IQR) <sup>a</sup> | 76 (68.3–86.5) | 76 (69–86) | 76 (68–86) | 0.39 |
| Level of education, N (%) <sup>c</sup> |  |  |  |  |
| None | 1 (0.5) | 0 (0) | 1 (1.0) | 0.98 |
| Primary | 28 (13.3) | 14 (13.6) | 14 (13.1) | 0.91 |
| Secondary | 136 (64.8) | 66 (64.1) | 70 (65.4) | 0.84 |
| University | 45 (21.4) | 23 (22.3) | 22 (20.7) | 0.76 |
| Marital status, N (%) <sup>c</sup> |  |  |  |  |
| Single | 53 (25.2) | 20 (19.4) | 33 (30.8) | 0.71 |
| Married | 32 (15.2) | 15 (14.6) | 17 (15.9) | 0.79 |
| Cohabitation | 124 (59.0) | 68 (66.0) | 56 (52.3) | 0.04 |
| Divorced | 1 (0.5) | 0 (0.0) | 1 (1.0) | 0.98 |
| Profession (Employed), N (%) <sup>c</sup> | 95 (45.2) | 45 (43.7) | 50 (46.7) | 0.66 |
| Monthly income (FCFA), median (IQR)<sup>a</sup> | 100K (70K–110K) | 100K (60K–100K) | 100K (71.3K–120K) | 0.27 |
| **Clinical parameters** |  |  |  |  |
| HBV positive, N (%) <sup>c</sup> | 7 (3.3) | 3 (2.9) | 4 (3.7) | 0.74 |
| Hemoglobin (g/dL), mean (SD) <sup>b</sup> | 11.10 | 11.3 (±1.3) | 10.9 (±1.1) | 0.04 |
| Multigravida, N (%) <sup>c</sup> | 176 (83.8) | 78 (75.7) | 98 (91.6) | 0.002 |
| Parity, N (%) <sup>c</sup> |  |  |  |  |
| Nullipara | 37 (17.6) | 23 (23.3) | 14 (13.1) | 0.06 |
| Primipara | 39 (18.6) | 16 (15.5) | 23 (21.5) | 0.27 |
| Multipara | 129 (61.4) | 59 (57.3) | 70 (65.4) | 0.23 |
| Missing | 4 (1.9) | 4 (3.9) | 0 (0) |  |
| ART regimen, N (%) |  |  |  |  |
| TDF/3TC/DTG | / | NA | 6 (5.6) |  |
| TDF/3TC/EFV | / | NA | 76 (71.0) |  |
| Other | / | NA | 2 (1.9) |  |
| Missing | / | NA | 23 (21.5) |  |
| Duration on ART (months), median (IQR)<sup>a</sup> | / | NA | 70 (23.3–103.8) |  |
| Cotrimoxazole prophylaxis, N (%) | / | NA | 44 (41.0) |  |
| **Birth parameters** |  |  |  |  |
| **Infant sex N (%)<sup>c</sup>** |  |  |  | 0.78 |
| Female | 113 (53.1) | 56 (54.3) | 57 (51.8) |  |
| Male | 100 (46.9) | 47 (45.7) | 53 (48.2) |  |
| **Term of delivery, N (%)<sup>c</sup>** |  |  |  |  |
| Preterm (<37 weeks) | 0 (0) | 0 (0) | 0 (0) | / |
| Term (37–42) | 198 (93) | 94 (91.3) | 104 (94.6) | 0.43 |
| Post-term (>42) | 0 (0) | 0 (0) | 0 (0) | / |
| Missing | 15 (7) | 9 (8.7) | 6 (5.4) |  |
| **Mode of delivery, N (%)<sup>c</sup>** |  |  |  |  |
| Per vaginal | 183 (92.4) | 89 (94.7) | 94 (90.5) | 0.29 |
| C-section | 10 (5.1) | 3 (3.2) | 7 (6.7) | 0.34 |
| Missing | 5 (2.5) | 2 (2.1) | 3 (2.8) |  |
| **Birth weight (g), median (IQR)<sup>a</sup>** | 3250 (3060-3515) | 3295 (3100-3538) | 3240 (3035-3515) | 0.41 |
| Normal (>2500 g), N (%)<sup>c</sup> | 181 (91.4) | 87 (92.6) | 94 (90.5) | 0.62 |
| Low (1500–2500 g) | 12 (6.0) | 5 (5.3) | 7 (6.7) | 0.77 |
| Missing | 5 (2.6) | 2 (2.1) | 3 (2.8) |  |
| **Birth length (cm), mean (SD)** | 50.4 (±2.6) | 50.6 (±2.5) | 50.3 (±2.8) | 0.49 |
| **Head circumference (cm), median (IQR)<sup>a</sup>** | 35.0, (34-36) | 35.0, (34-36) | 36.0, (34-36) | 0.05 |
| **Infant feeding (first 6 months), N (%)<sup>c</sup>** |  |  |  |  |
| Exclusive breastfeeding | 46 (29.6) | 19 (29.3) | 27 (30.0) | >0.99 |
| Formula feeding | 35 (22.6) | 1 (1.5) | 34 (37.8) | <0.0001 |
| Mixed feeding | 53 (34.2) | 31 (47.7) | 22 (24.4) | 0.004 |
| Missing | 21 (13.6) | 14 (21.5) | 7 (7.8) |  |

<sup>a</sup> = Mann-Whitney test; <sup>b</sup> = Unpaired t test, <sup>c</sup> = Chi Square test, K = × 1000

Birth outcomes (term of delivery, mode of delivery, birth weight, birth length and head circumference) were similar between HUU (94) and HEU (103) **Table 1**. Most infants in both groups HUU (91.3%) and HEU (94.6%) were born at term by vaginal delivery (HUU = 94.7%, HEU= 90.5%) and had a normal weight (HUU 92.6% and HEU= 90.5%). Median birth weight was 3295g; IQR=3100g-3538g and 3240; IQR=3035g-3515g in both HUU and HEU groups respectively. During the first six months of life, the proportion of HUU and HEU infants who were exclusively breastfed were similar (29.3% and 30.0%; ˃0.99 for HUU and HEU respectively). This was not similar to mixed feeding (47.7% and 24.4%; P=0.004 for HUU and HEU respectively). Nevertheless, the proportion of HEU on formula feeding was higher than HUU (1.5% and 37.8% for HUU and HEU respectively; P=<0.0001) with 61.7% (21/34) of the formula fed HEU being male infants **Table 1**.

### Growth outcomes in HEU vs HUU

Baseline sex stratified comparisons of HEU and HUU infants WHZ, HAZ, WAZ, BAZ and HCZ Z-scores across 4 timepoints (months 3, 6, 9, and 12) revealed at month 3, that male HEU experienced low WAZ (1.07 ±1.94 vs 0.05 ±1.35; P=0.04 for HUU and HEU respectively) and low HCZ (1.39 ±2.38 vs 0.43 ±1.17; P=0.03 for HUU and HEU respectively) compared to their HUU peers. No other sex-based differences were observed in HEU vs HUU **Supplementary table 3**. When analysis was restricted to paired samples in the cohort, the difference in WAZ observed among males at month 3 persisted (P=0.04) **Figure 2a**. Infants’ nutritional classification by WHO criteria were similar except for stunting which was more prevalent in female HEU at month 6 (0% vs 21%; P=0.01) and among male HEU at month 3 (0% vs 17.4%; P=0.01) and month 12 (12.5% vs 38.9%; P=0.03) compared to their HUU counterparts’ **Supplementary table 4**. In males the difference observed at month 6 persisted to month 12 (P=0.04) **Fig 2b**. To ascertain if these differences were driven by maternal HIV, mixed-effects regression models were fitted, adjusting for socioeconomic status (SES), maternal and gestational age, while accounting for repeated measures (3, 6, 9, and 12 months). In females, adjusted analysis showed lower WAZ (β –0.84; 95% CI –1.74, 0.07; P=0.071) and HAZ (β –1.13 95% CI –2.29, 0.03; P=0.057) in HEU compared to HUU. Male HEU had lower HAZ both in the unadjusted (β –0.73; 95% CI –1.44, –0.02; P=0.043) and adjusted models (β –0.75; 95% CI –1.49, –0.01; P=0.047) highlighting the difference in linear growth for children born to mothers with HIV **Supplementary table 5**. Overall, HEU exhibit sex-specific growth vulnerabilities with more consistent linear growth deficits observed in males during the first year of life.

**Figure 2.**
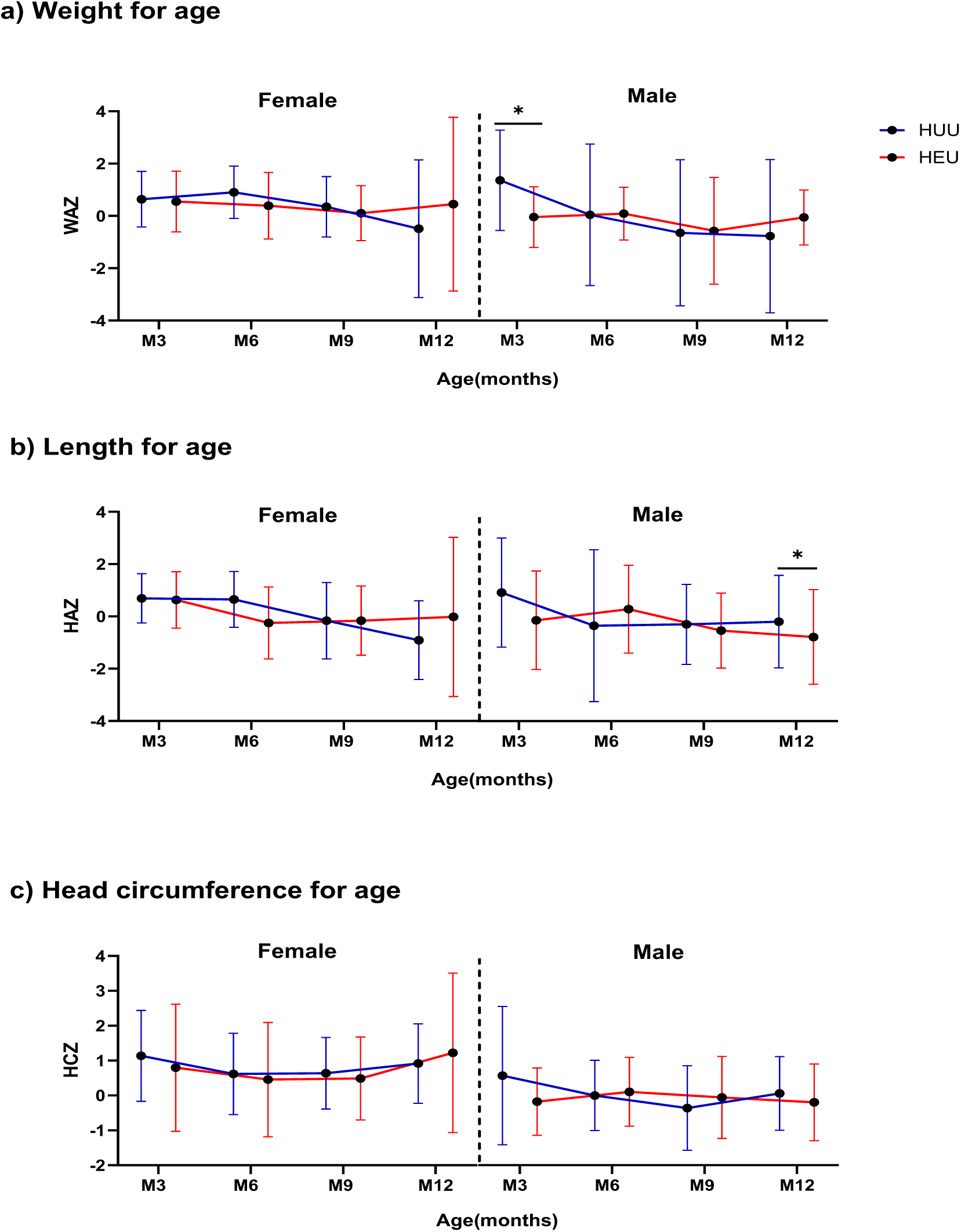
Growth curves for **a)** Weight, **b)** Length and **c)** Head circumference for age measured at 3, 6, 9 and 12 months of age in female and male HUU and HEU infants plotted as means and standard deviation using z-scores. HUU: Female N=18, Male N=17. HEU: Female N=30, Male N=28. *Note: The dependent t test was used for normally distributed data while the Wilcoxon matched-pairs signed rank test was used to compare non-normally distributed data sets*.

### Sex-stratified differences in symptom prevalence among HEU and HUU

Irrespective of the groups, the prevalence of self-reported illness symptoms increased from 6 weeks to 12 months of age **Figure 3a**. HEU were twice more likely to have rhinorrhea-related symptom than HUU at 9 months of age (RR=2.00; 95% CI 1.23, 3.26; P=0.005). HEU had a 0.53 lower RR of acquiring a fever (95% CI 0.32, 0.85; P=0.01) and a 0.59 RR of developing a cough (95% CI 0.37, 0.93; P=0.03) than their HUU counterparts at month 12 **Figure 3b**. HEU males but not females had more rhinorrhea symptoms than HUU counterparts at month 9 (RR=2.29; 95% CI 1.17, 4.79; P=0.02) whereas at month 12 the HEU female but not male showed lower prevalence of reported fever (RR=0.31; 95% CI 0.15, 0.63; P=0.002) and cough (RR=0.23; 0.23; 95% CI 0.08, 0.58; P=0.002) compared to the female HUU **Figure 3c**. Overall, illness symptoms increase with age in both groups with male HEU displaying higher rhinorrhea at 9 months.

**Figure 3:**
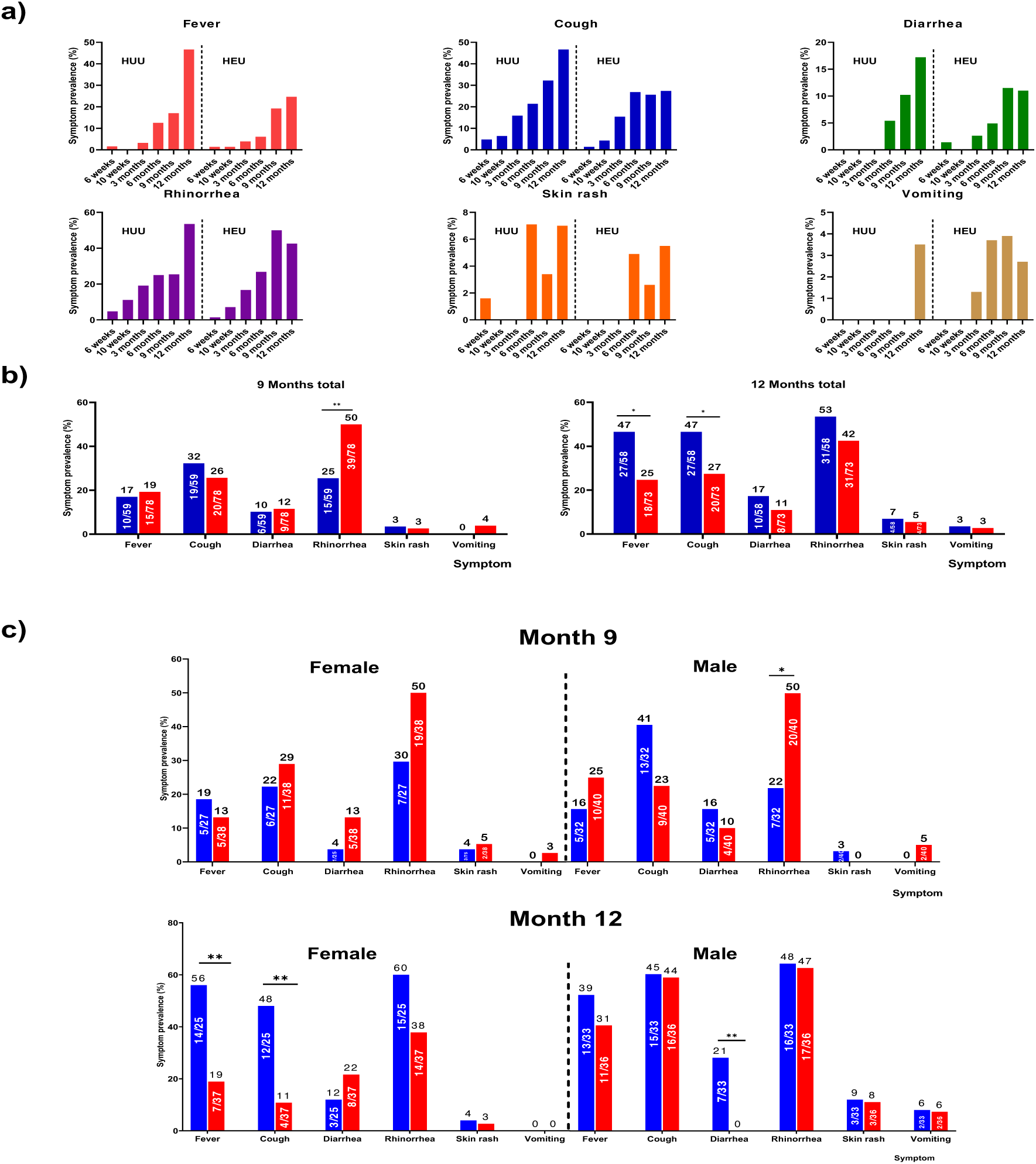
Longitudinal trend and prevalence of maternal self-reported Symptoms (fever, cough, diarrhea, rhinorrhea, skin rash, and vomiting) in HEU (red) and HUU (blue) Infants during the First Year of Life. **a)** Increase in prevalence of symptoms from 6 weeks to 12 months. **b)** HEU experience higher prevalence of rhinorrhea at month 9. **c)** Male HEU experience higher rates of rhinorrhea at month 9. ***Note:*** *Pearson’s Chi-square/Fisher’s exact test was used for categorical data to determine the differences in symptom rates in HEU and HUU infants*.

### Prevalence and viremia of infections in HEU and HUU stratified by HIV exposure and sex

To further assess the burden of infections in our study groups, the prevalence and viremia of CMV, rotavirus, malaria, rhinovirus, RSV, Influenza A & B were determined from 3 to 12 months of age in HEU and HUU infants. While infections with rotavirus, malaria, RSV, and Influenza A & B were quasi-inexistent in both groups from month 3 to 12 with a prevalence ranging from 1.4% to 5.5%, CMV and Rhinovirus infections were ubiquitous from 3 to 12 months with prevalence ranging from 33.9% to 60.7% for CMV and 25.3% to 50.0% for Rhinovirus infections by month 12 **Supplementary figure 1**. At month 6, CMV prevalence was higher in the HUU compared to the HEU (60.7% vs 43.2%, P=0.03), with no other differences in prevalence amongst HEU and HUU. Sex stratified analysis of CMV, and rhinovirus infection prevalence showed no differences between male HEU and HUU, or female HEU and HUU across all timepoints **Figure 4a and b**. Sex stratified analysis of CMV and Rhinovirus viremia among PCR positives showed lower viremia in the female HEU compared to their HUU counterparts at month 9 (P=0.013) **Figure 4 c and d**. Across the first year of life, infection burden among HEU and HUU were largely comparable with CMV with rhinovirus being the most frequent infection.

**Figure 4:**
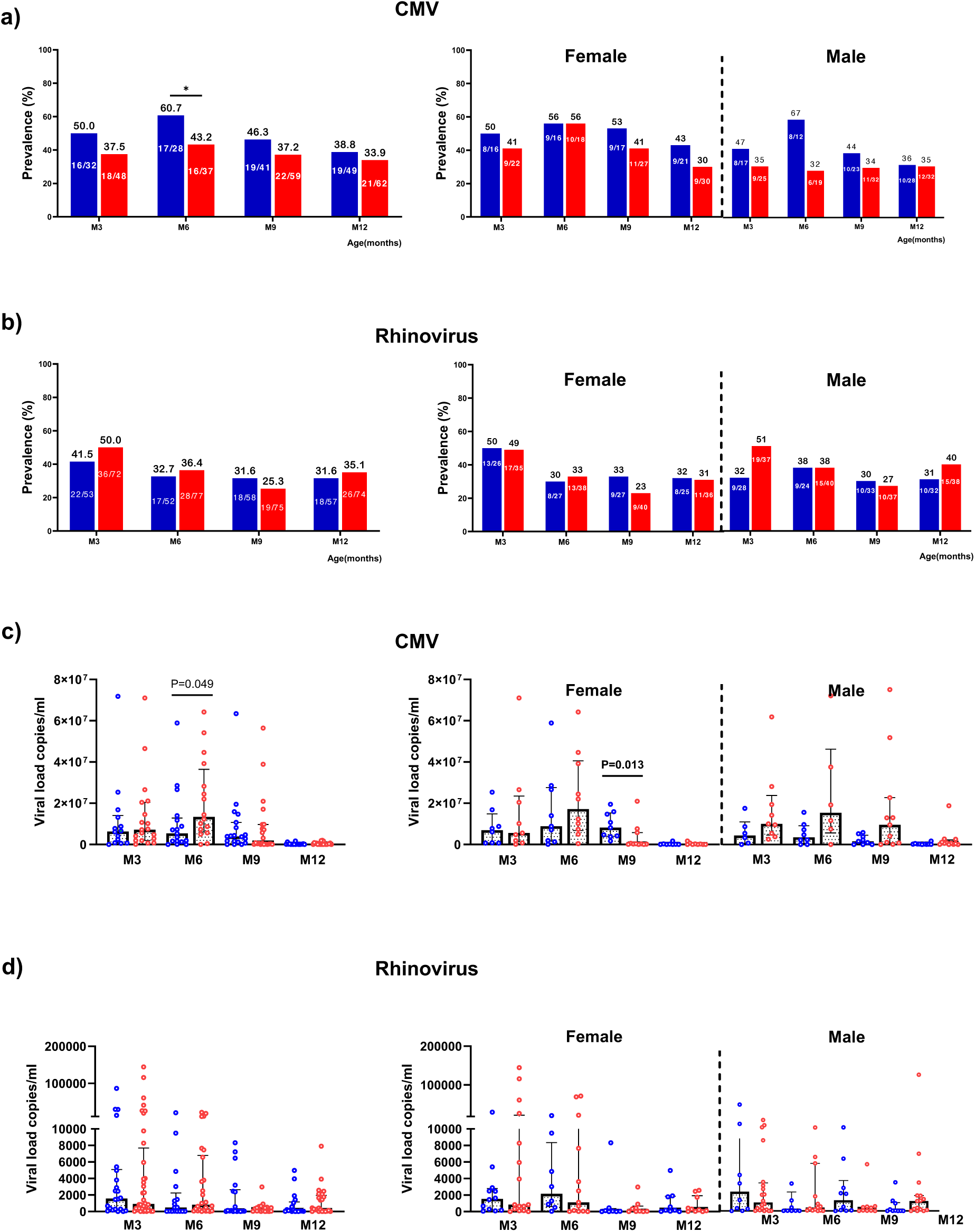
Prevalence and viremia of CMV and Rhinovirus infections in HEU (**red**) and HUU (**blue**) at 3, 6, 9 and 12 months of age. **a)** HIV unexposed uninfected infants (HUU) experience a higher prevalence of CMV at 6 months of age with no sex-based differences. **b)** Comparable prevalence of Rhinovirus infections amongst HEU and HUU infants with no sex-based differences observed. **c)** Higher CMV viremia in HEU infants at 6 months of age. Female HUU infants experience a higher viremia at 9 months of age **d)** Comparable Rhinovirus viremia in HEU and HUU infants with no sex-based differences. Note: *Pearson’s Chi-square test was used to determine the differences in HEU and HUU infants and independent t test was used for normally distributed data while the Mann Whitney test was used to compare non-normally distributed data sets*.

### Antibody transfer and dynamics in HEU and HUU

Following our observation of sex-effects on self-reported illness symptom patterns and PCR-based detection of common childhood infections, we evaluated humoral immune response to 3 pathogens, CMV, RSV and enterovirus to explore whether altered antibody profiles align with sex-related disparities of infection rates. Levels of antigen-specific IgG were measured at 5 time points, (birth, month 3, 6, 9 and 12). CMV, antibody levels peaked at birth and decreased with time in both groups and sexes. Female HEU had higher titers at month 3 compared to the HUU females (P=0.04) **Figure 5a**. Maternal-infants antibody titer correlations suggest all infants lost maternal CMV antibodies by month 9 **Supplementary figure 2a**. RSV antibody levels also peaked at birth and diminished steadily till month 9 and increased by month 12 in both groups and sexes with no difference by exposure and sex **Figure 5b**. Maternal-infant antibody level correlation showed that maternal antibodies were minimal by month 6 **Supplementary figure 2b.** Enterovirus antibody levels were highest at birth and diminished to their lowest levels at 6 months, then started rising at months 9 with no significant differences between HEU and HUU. At birth, the male HEU infants had lower titers compared to their HUU counterparts (P=0.0066) **Figure 5c**. Put together, these findings indicate that maternally derived protection against CMV; RSV and enterovirus declined similarly in HEU and HUU, suggesting that altered humoral transfer is unlikely to completely account for the sex specific disparities observed in infection rates.

**Figure 5.**
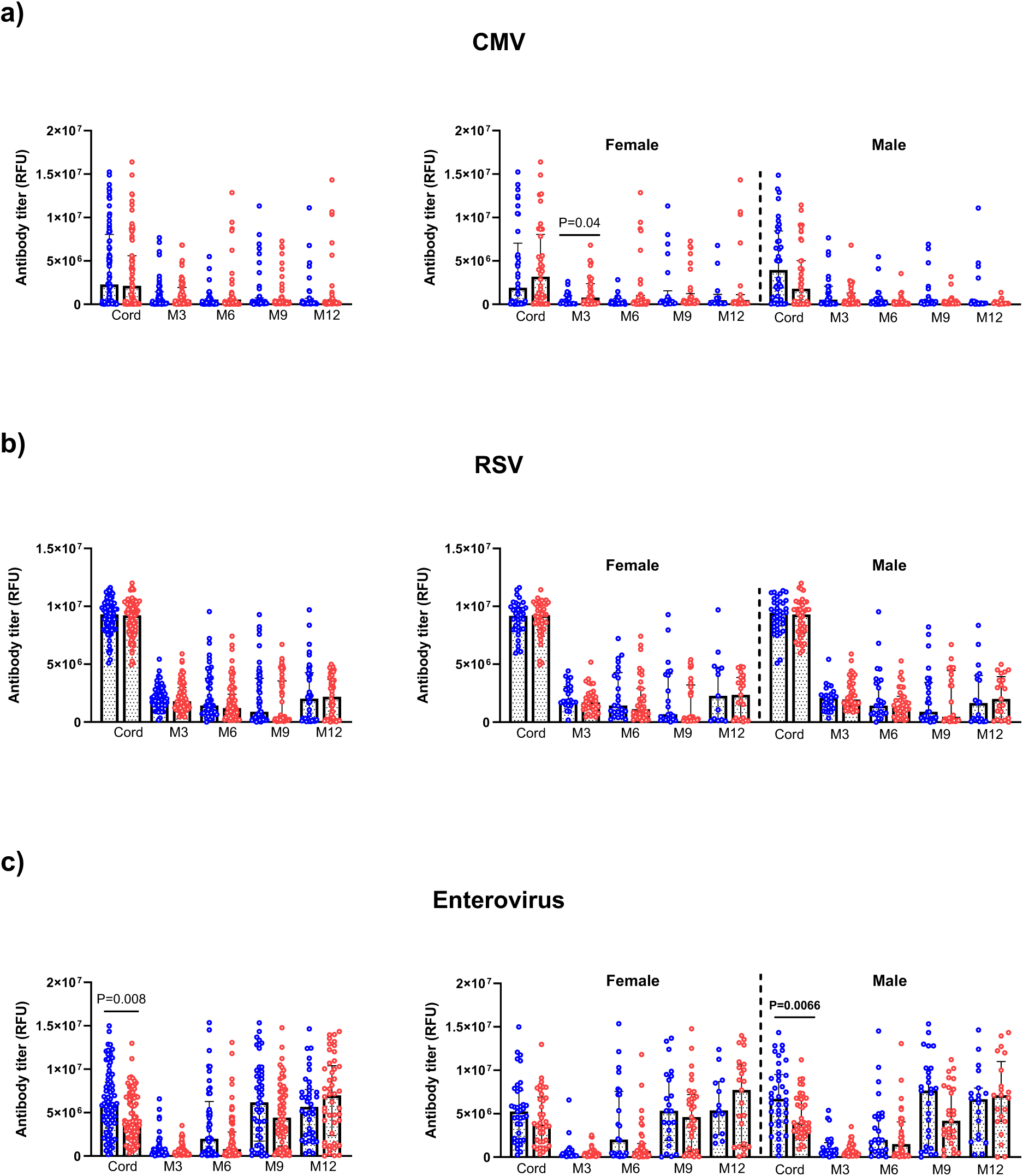
Sex stratified comparative longitudinal analysis of antibody levels to **a)** CMV, **b)** RSV and **c)** Enterovirus from birth till 12 months of age in HEU (**red**) against their HUU (**blue**) counterparts. *Note: For comparative analysis, the independent t test was used for normally distributed data sets and the Mann-Whitney test for data that failed to pass normality test*.

Antibody trends described are done on independent samples. Sample sizes for each comparison= Female; cord (HUU=37 HEU=38), M3 (HUU=23 HEU=34), M6 (HUU=24 HEU=37), M9 (HUU=22 HEU=33), M12 (HUU=13 HEU=23) and Male; cord (HUU=38 HEU=40), M3 (HUU=27 HEU=36), M6 (HUU=25 HEU=36), M9 (HUU=27 HEU=27), M12 (HUU=19 HEU=21).

## Discussion

This study explored early-life growth trajectories, clinical trends associated with infection dynamics and humoral immune responses among HEU and HUU infants during their first year of life. Globally, our data suggest that HEU infants showed growth and infection profiles that was largely like that of their HUU peers, with a few transient, sex-specific differences in growth, infection dynamics and antibody responses.

The feeding practices observed in this cohort is reminiscent of what has been reported across PMTCT studies, with higher formula use amongst the HEU but similar exclusive breastfeeding rates, where HIV-exposed mothers are more likely to practice mixed feeding due to perceived risk of transmission [20,21]. Despite similar breastfeeding rates, HEU infants exhibited sex-specific growth differences with the female HEU experiencing increased stunting at 6 months while male infants portrayed lower WAZ and HCZ at 3 months and persistent lower HAZ at 12 months. These findings are consistent with many reports from several studies showing that stunting is the most observed growth defect amongst HEU infants even when adjusted for confounders such as socioeconomic status[22–24]. Some other studies argue that birth characteristics such as sex are predictive in growth trajectories, hence emphasizing the need to account for sex in growth analysis[25,26]. Our findings show that despite the widespread use of ART, in utero exposure to HIV subtly affects early growth, with short-term outcomes differing by sex.

The temporal trends of symptoms reported by caregivers and the detection of pathogens in our study sheds light on key features of infection dynamics in infancy. The rise in the prevalence of symptoms from 6 weeks to 12 months of age aligns with growing social contact, fading maternal antibodies, and increasing exposure to respiratory and enteric pathogens. Cough and rhinorrhea were the most frequent symptoms in both groups; this observation reflects the dominance of respiratory infections during infancy[27,28]. Interestingly, at 9 months, HEU infants exhibited higher rhinorrhea prevalence compared to HUU, this observation was driven by the male HEU and suggests a possible increased susceptibility to respiratory infections at this age, more importantly in the males. This finding aligns with the evidence that early life immunity is shaped by sex hormones and genetic factors, with females often portraying a more robust antiviral response [29,30]. These differences could also reflect variations in maternal antibody transfer, immune activation and postnatal exposure between sexes. This appeared to be transient, as the tendency was reversed at month 12 with a lower prevalence of fever and cough. The later suggests a more efficient immune control or reduced exposure given that HIV-affected families are prone to protective behaviors[31].

During the last decades, several studies showed that HEU had increased mortality and morbidity rates when compared to HUU and this was mainly associated to respiratory infections as suggested by symptoms rates described in this study[2,5–8,32]. In this cohort, CMV and Rhinovirus emerged as the predominant infections, consistent with their omnipresence in early infancy[6,33,34]. The comparable infection rates between HEU and HUU suggest similar exposure risks, although higher CMV viremia at 6 months in HEU might indicate temporarily reduced viral containment or delayed immune maturation. Similar exposure risks and effective viral control hypothesis are further strengthened by the quasi-existence of other infections screened in this study. This finding is consistent with studies that reported no significant difference in infection prevalence of CMV, RSV and asymptomatic malaria between HEU and HUU potentially be attributed to the effective implementation of PMTCT programs [23,35,36]. Despite similarity in infection prevalence across groups, HEU infants remain prone to severe infection outcomes[2,6,37,38], underscoring the need to emphasize studies of the vulnerability of HEU on disease severity.

Patterns of humoral immune responses paralleled the infection data. Antibody levels for CMV, followed the expected maternal decay curve with no differences between groups except for momentarily higher titers in the female HEU group at 3 months of age. This suggests an efficient transfer of maternal antibodies and subsequent retention in the HEU infants with the female HEU showing a greater retention rate by month 3 compared to their HUU female peers. The enhanced retention may be influenced by immune activation and the higher susceptibility of HIV infected mothers to CMV reactivation[39,40]. On the contrary, RSV and enterovirus antibody profiles showed rapid decay of antibodies by months 3 and 6 followed by a gradual recovery due to postnatal exposure. The lack of difference between HEU and HUU shows that HIV exposure did not significantly impair antibody production with the first year of life.

Put together, these findings illustrate a complex interplay between HIV exposure, sex, growth and pathogen specific immunity. This underscores the fact that, while maternal HIV exposure might not consistently affect infant growth or immunity, it can interact transiently with biological sex to shape growth and infection rates in HEU infants. The transient nature of differences observed, supports the hypothesis that immune maturation compensates for early disparities over time.

We acknowledge several shortcomings in this study. First, the symptoms were self-reported by caregivers, this may introduce subjective bias. Secondly, despite 1014 infants timepoints from birth through months 12, our study may be underpowered for the detection of subtle changes in infection across subgroups. Finally, while antibody profiles provide a primary understanding of immune responses, cellular immune correlates would provide complementary insight.

In sum, this study offers an integrated analysis of infection dynamics, growth, and antibody responses among HEU and HUU, shedding light on comparable overall outcomes with modest transient sex-based differences, emphasizing the importance of sex as a biological variable in pediatric infection and immunity research. Upcoming work should integrate cellular immune phenotyping and longer-term-follow-up to elucidate the developmental and immunological trajectories of HEU.

## Data Availability

All data produced in the present study are available upon reasonable request to the authors

## Author contribution

FEL, KN and MB contributed to conceptualization. HA, KT and MT were involved in data collection; RD, RN, MS, EG were involved in laboratory assays; RD, RN, MS, EG MB performed data analysis; KN, MB and FEL were involved in Validation; KN, FEL and MB were involved in funding acquisition, FEL, MB, KN, CK, JB, RL were involved in project administration, CK, JB, RL supervised the study; RD wrote the original draft and all authors were involved in reviewing and editing.

## Acknowledgments

We acknowledge the mothers and children who participated in this study, as well as the staff of the HIV and Maternity Units of CASS Nkoldongo. We also sincerely thank Djeukeng Christela, Hillary Tene, and Daniela Ebamu for their invaluable support with specimen processing during periods of high workload.

## Supplementary Materials

**Supplementary table 1:**
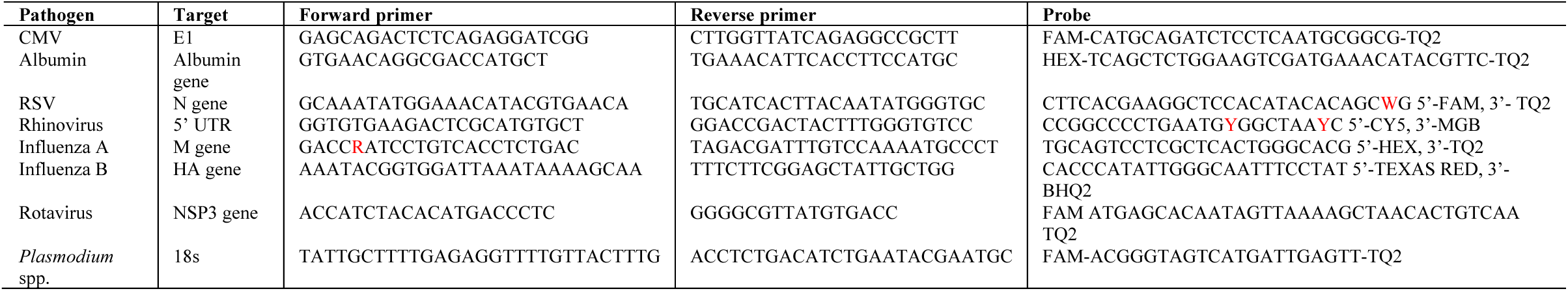
Primer and probe sequences used for multiplex RT-PCR detection of pathogens.

**Supplementary table 2:** Cycling conditions.

| Pathogen panel | Reverse transcription | Initial denaturation | Cycling steps (Den, Ann, Elon) | Total cycles |
| --- | --- | --- | --- | --- |
| Multiplex DNA (CMV + Albumin) | / | 95°C 2min | 94°C 40s, 57°C 30s, 72°C 1min | 41 |
| Multiplex RNA (RSV, Rhino, Influenza A & B) | 55 °C 10min | 95 °C 5min | 94°C 15s, 60°C 20s, 72°C 40s | 42 |
| Rotavirus (singleplex) | 55 °C 10min | 95 °C 5min | 94°C 10s, 60°C 20s, 60°C 40s | 40 |
| Malaria (singleplex) | / | 95°C 10min | 95°C 15s, 58°C 20s, 58°C 40s | 47 |

**Supplementary Table 3:**
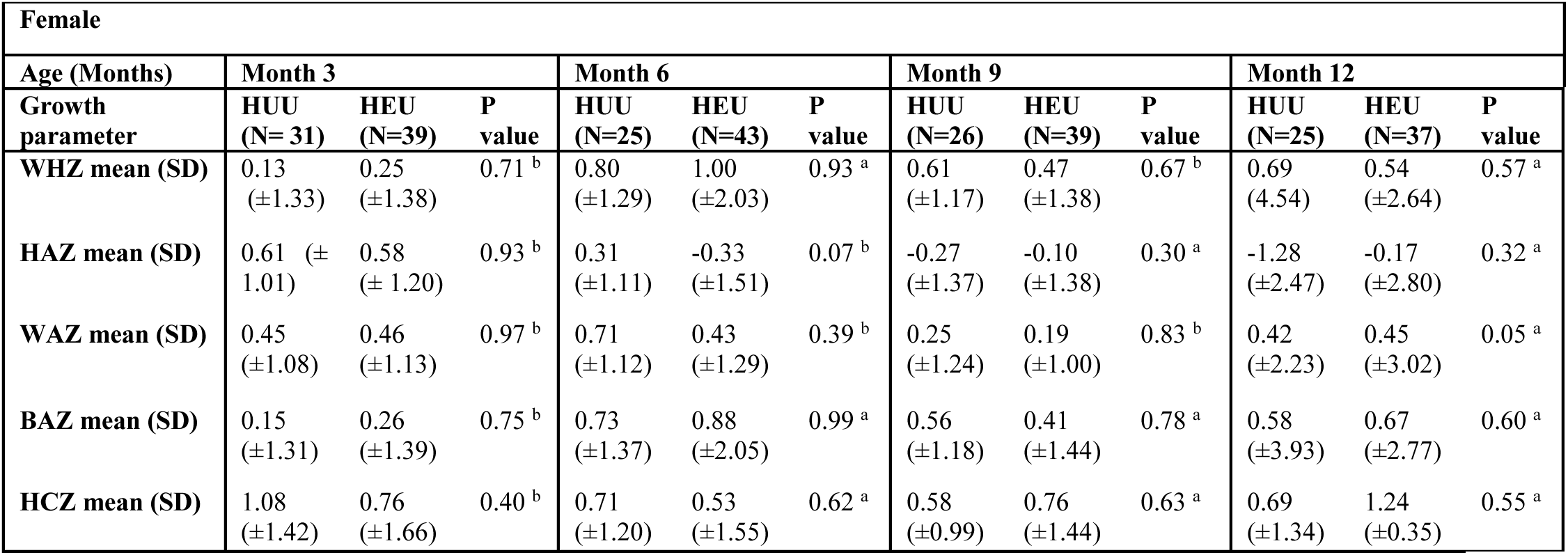
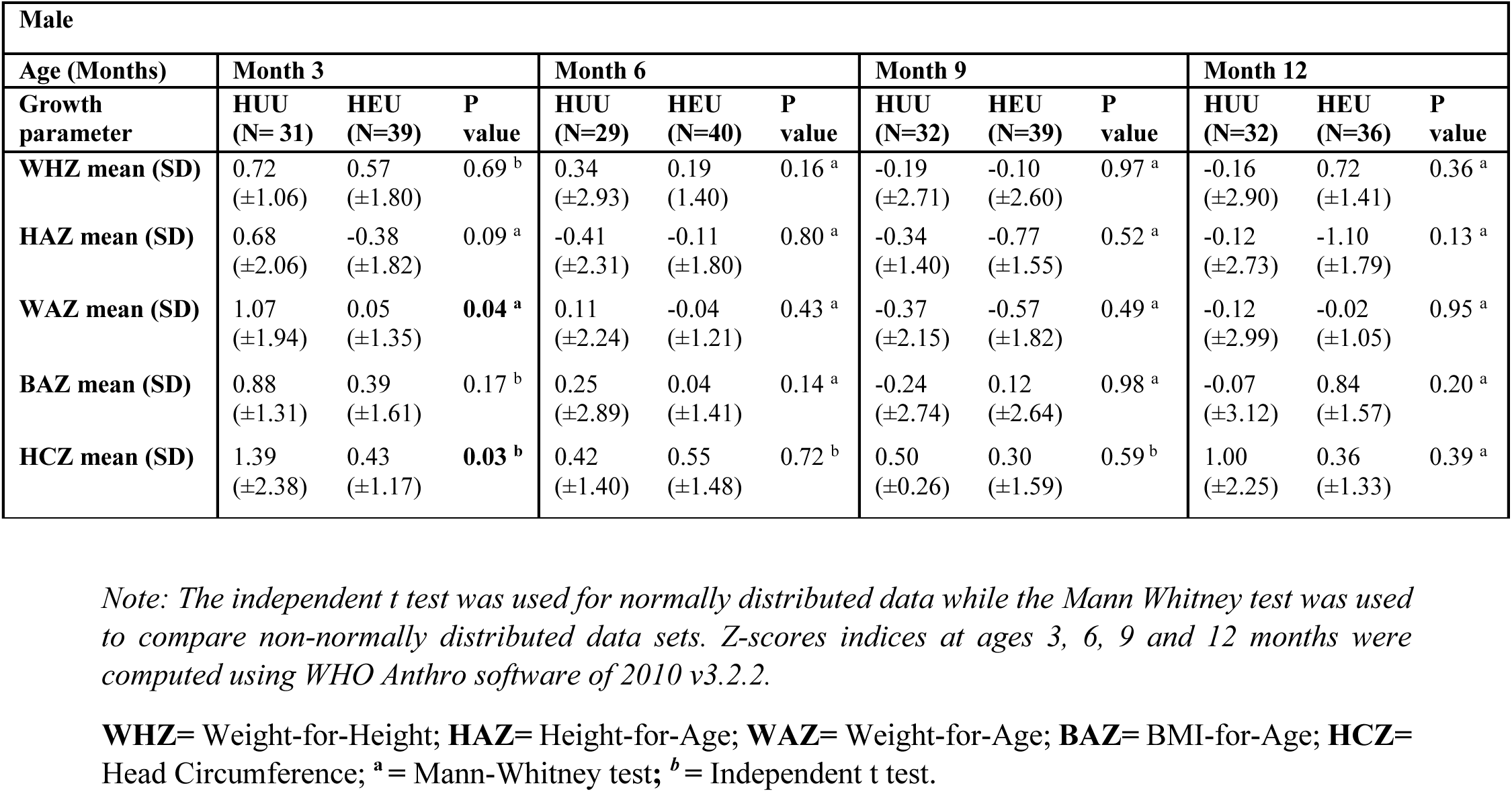
Sex stratified Z-score indices of infants at 3, 6, 9 and 12 months of age by HIV exposure status.

**Supplementary table 4:** Sex stratified nutritional classification of infants at 3,6,9 and 12 months.

| Female |  |  |  |  |  |  |  |  |  |  |  |  |
| --- | --- | --- | --- | --- | --- | --- | --- | --- | --- | --- | --- | --- |
| Age (Months) | Month 3 |  |  | Month6 |  |  | Month 9 |  |  | Month 12 |  |  |
| Nutritional classification | HUU (N= 30) | HEU (N=39) | P value | HUU (N=26) | HEU (N=42) | P value | HUU (N=26) | HEU (N=39) | P value | HUU (N=25) | HEU (N=36) | P value |
| Wasting, n (%)<sup>c</sup> | 1(3.3) | 2(5.1) | >0.99 | 0(0) | 1(2.3) | >0.99 | 0(0) | 2(5.1) | 0.51 | 2(8) | 1(2.8) | 0.56 |
| Stunting, n (%)<sup>c</sup> | 1(3.3) | 1(2.6) | >0.99 | 0(0) | 9(21.4) | **0.01** | 2(7.7) | 2(5.1) | >0.99 | 8(32) | 6(16.7) | 0.22 |
| Underweight, n (%)<sup>c</sup> | 2(6.7) | 0(0) | 0.19 | 0(0) | 1(2.3) | >0.99 | 1(3.7) | 0(0) | 0.41 | 2(8) | 5(13.9) | 0.69 |
| Microcephaly, n (%)<sup>c</sup> | 1(3.3) | 2(5.1) | >0.99 | 0(0) | 1(2.3) | >0.99 | 0(0) | 1(2.6) | >0.99 | 1(4) | 0(0) | 0.41 |

| Male |  |  |  |  |  |  |  |  |  |  |  |  |
| --- | --- | --- | --- | --- | --- | --- | --- | --- | --- | --- | --- | --- |
| Age (Months) | Month 3 |  |  | Month6 |  |  | Month 9 |  |  | Month 12 |  |  |
| Nutritional classification | HUU (N= 31) | HEU (N=39) | P value | HUU (N=29) | HEU (N=40) | P value | HUU (N=32) | HEU (N=39) | P value | HUU (N=32) | HEU (N=36) | P value |
| Wasting, n (%)<sup>c</sup> | 0(0) | 1(2.6) | >0.99 | 2(7.1) | 2(5) | >0.99 | 2(6.3) | 4(103) | 0.68 | 4(12.5) | 2(8.3) | 0.69 |
| **Stunting, n (%)<sup>c</sup>** | 0(0) | 7(18.4) | **0.01** | 6(21.4) | 8(20.0) | >0.99 | 3(9.4) | 9(23.1) | 0.20 | 4(12.5) | 14(38.9) | **0.03** |
| **Underweight, n (%)<sup>c</sup>** | 1(3.2) | 2(5.1) | 0.62 | 1(3.6) | 2(5) | >0.99 | 3(9.4) | 2(5.1) | 0.65 | 4(12.5) | 1(2.9) | 0.18 |
| **Microcephaly, n (%)<sup>c</sup>** | 1(3.2) | 1(2.6) | >0.99 | 1(3.6) | 3(8.3) | 0.63 | 1(3.1) | 2(5.1) | >0.99 | 0(0) | 1(2.8) | >0.99 |

*Note: Pearson's Chi-square/Fisher's exact test was used for categorical data to determine the differences*
*in HEU and HUU infants.*
**Wasting** = WHZ < -2, **Stunting** = HAZ < -2, **Underweight** =WAZ < -2, **Microcephaly** =HCZ < -2, <sup>c</sup> =
Chi Square test

**Supplementary table 5:**
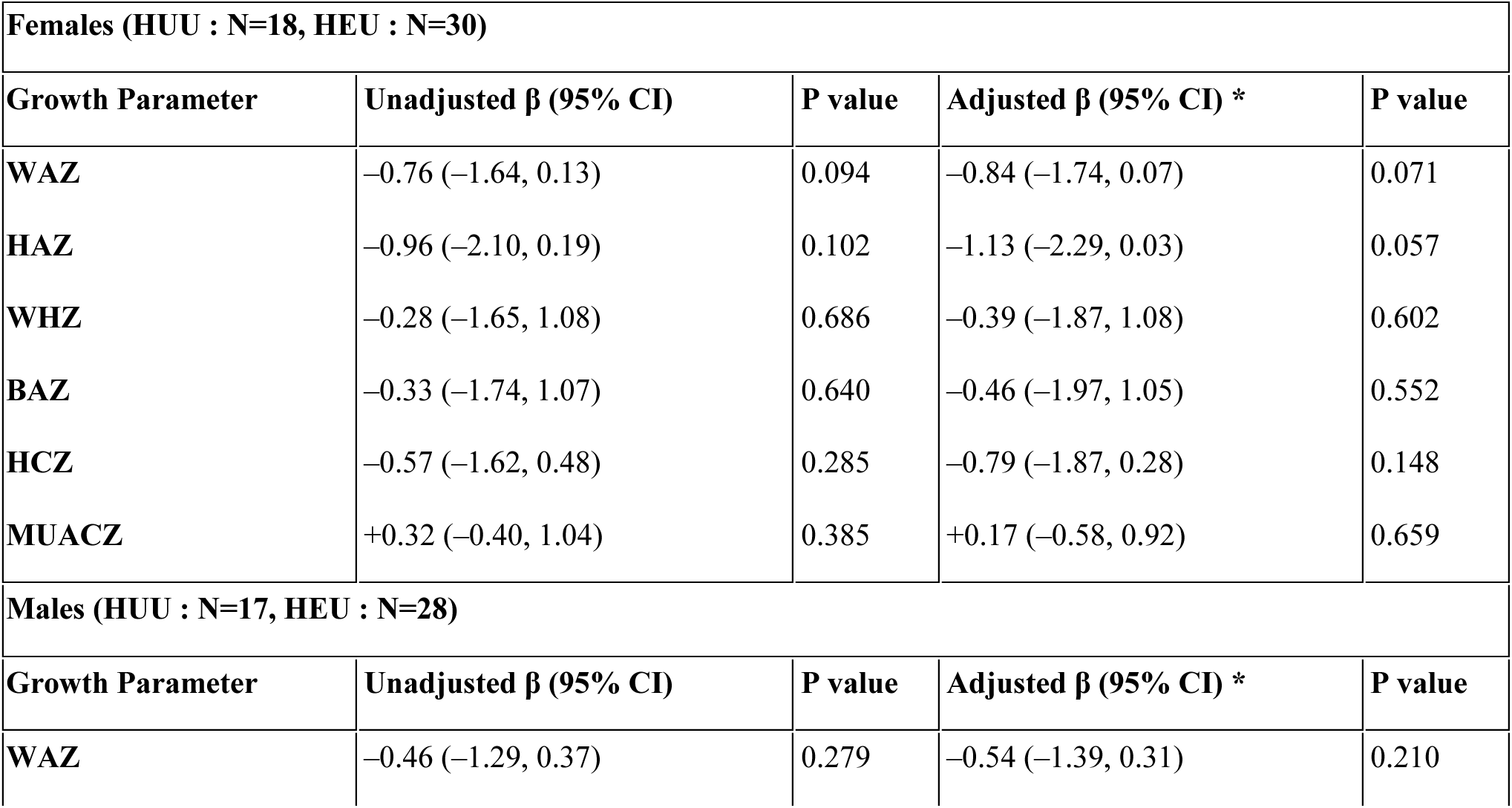
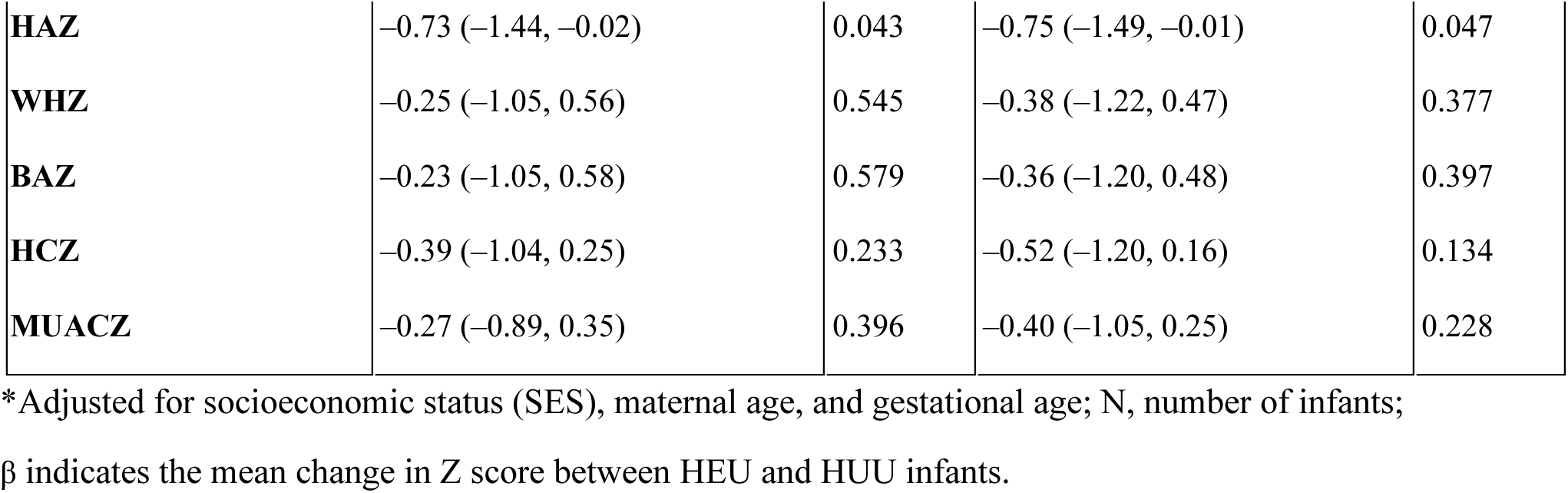
Mixed-Effects Regression for longitudinal comparisons of growth parameters between HEU and HUU infants.

**Supplementary figure 1:**
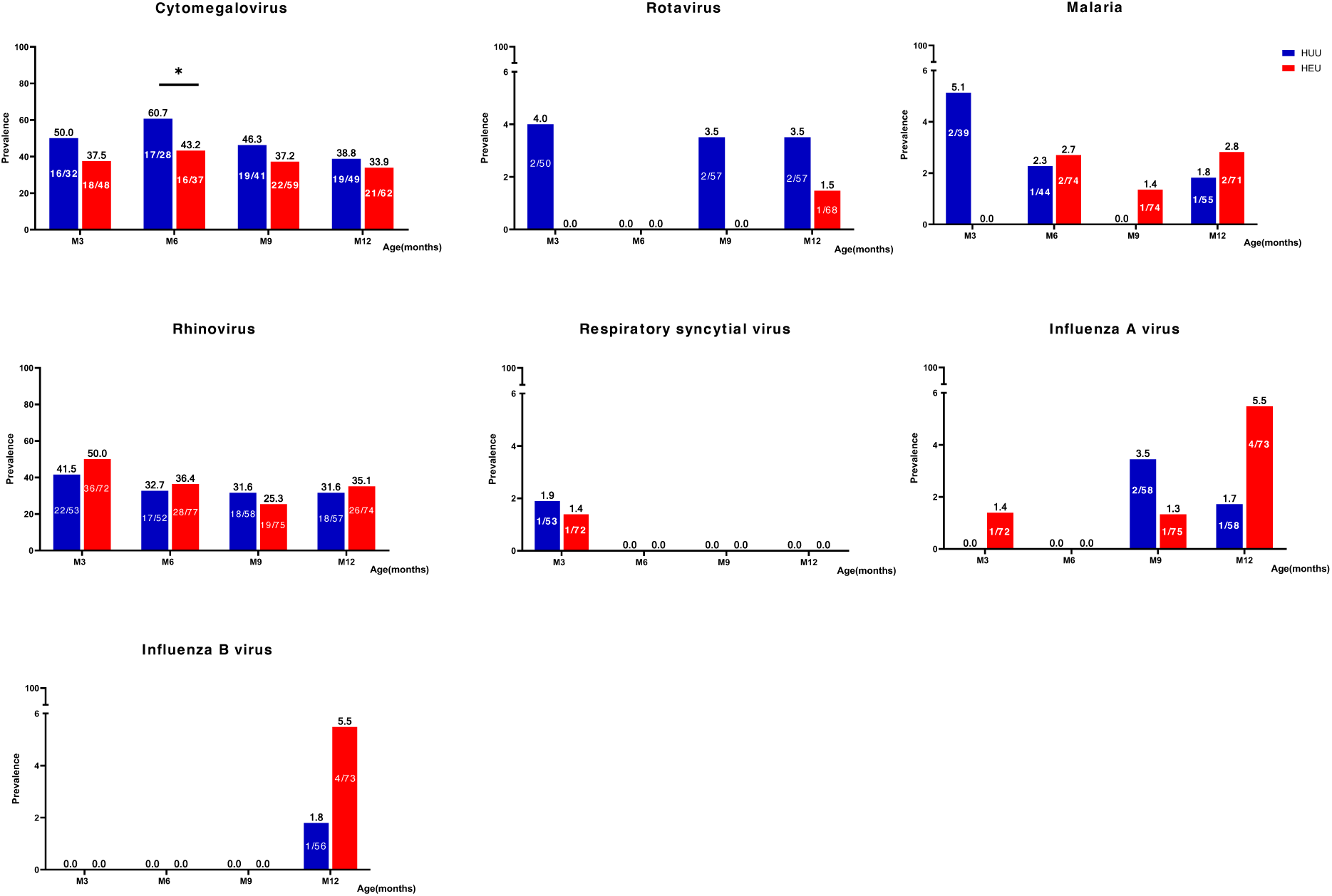
Prevalence of 7 common childhood infections by HIV exposure status from month 3 to 12. Note: *Pearson’s Chi-square test was used to determine the differences in HEU and HUU infants*.

**Supplementary figure 2:**
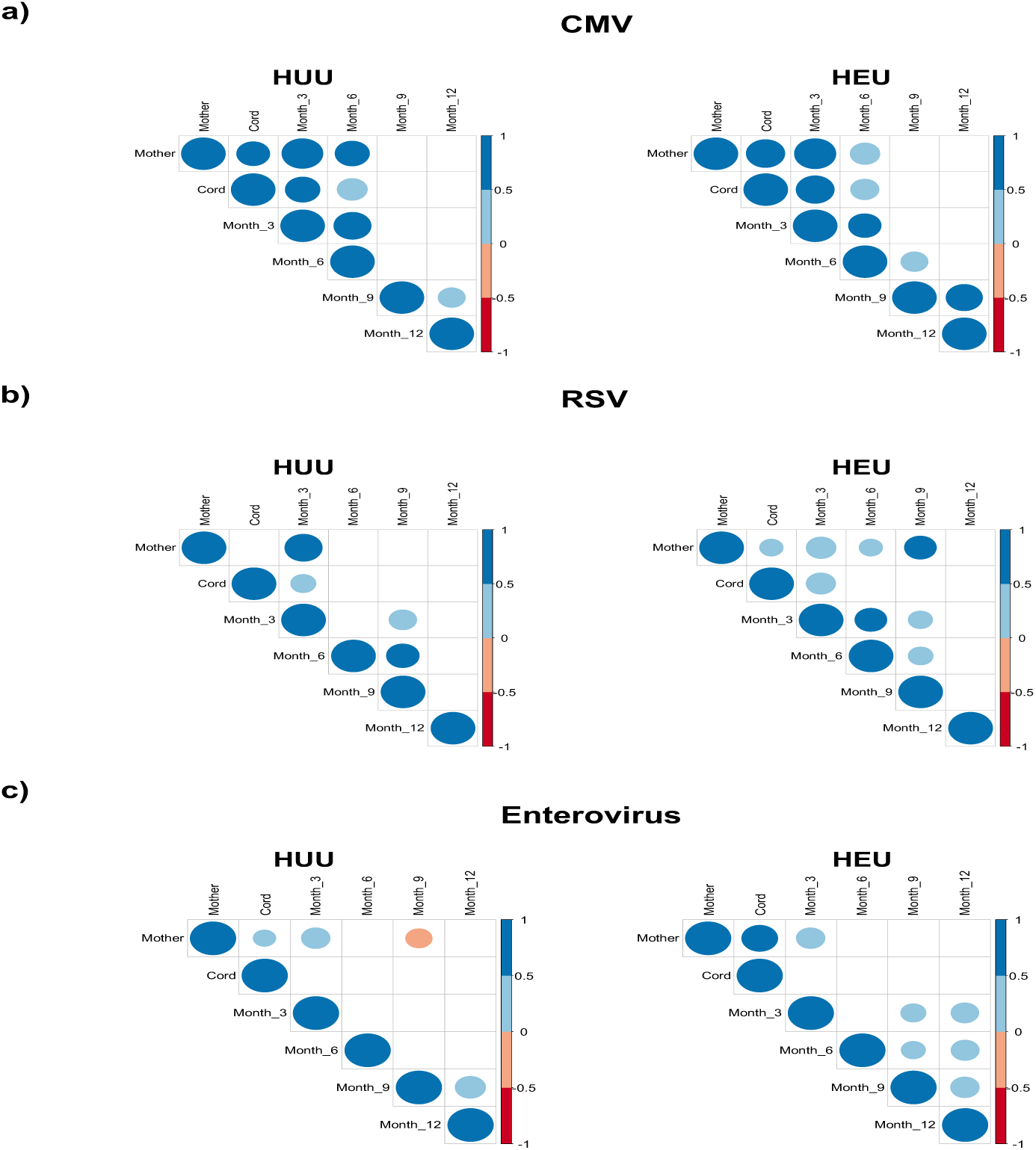
Correlation of maternal and cord antibodies to months 3, 6, 9 and 12 titers in HEU and HUU infants. *Note: Only correlations with p < 0.05 appear on correlograms. The size of the circle represents the correlation coefficient, with positive and negative values in blue and red shades respectively*.

